# A Deep Learning Framework for Causal Inference in Clinical Trial Design: The CURE AI Large Clinicogenomic Foundation Model

**DOI:** 10.1101/2025.03.06.25323534

**Authors:** Amit D Weiss, Vitalay Fomin, Di Feng, Zuojian Tang, James Cai, Bino John, Neil T Pfister

## Abstract

Clinical research is limited by the capability to define the most important combinations of clinical features and biomarkers that predict therapeutic benefit. Here, we introduce CURE AI (Clinical trials Uncovering Real Efficacy Artificial Intelligence), a novel deep learning framework designed to predict individual patient benefit from a new therapeutic intervention compared to a standard of care. CURE AI utilizes a large clinicogenomic foundation model to understand the complex relationships between the vast clinical and multiomic features in clinical trial data. To build CURE AI, we trained a proprietary foundation model based on a deep learning architecture and training schema using a large collection of clinical and multiomics datasets. Using CURE AI, we seek to understand the complex interplay between clinicogenomic information from clinical trial arms to predict the magnitude of therapeutic benefit on the individual patient level. In this paper, we finetuned the CURE AI foundation model on lung cancer data from the OAK non-small cell lung cancer clinical trial. We observed that the trial could have been significant for progression-free survival (PFS) with fewer than half of the patients enrolled using CURE AI to guide trial enrollment (p = 0.60 to p < 0.05). The finetuned CURE AI (termed CURE Lung Cancer) demonstrated direct generalizability on a held-out independent clinical trial dataset, the POPLAR trial, by converting an insignificant PFS endpoint to significance while also including the majority of patients (88%; p = 0.21 to p < 0.05). In summary, we developed a causally-aware clinicogenomic deep learning platform that can learn to predict individualized patient benefit of investigational therapy compared to an existing standard of care. Because we use a foundation model trained on readily measurable patient characteristics, CURE AI can be applied to a variety of scientific and clinical uses including adaptive clinical trials, toxicity prediction, treatment response prediction, and understanding of drug resistance and response mechanisms.

## Introduction

Clinical research is hampered by limitations in identifying the most important combinations of clinical and biologic features that can predict therapeutic response. New biomarker discovery is crucial to help define patient populations eligible for therapies. When large scale molecular profiling of patients (e.g., transcriptomic or proteomic) are carried out, as in OAK, POPLAR and other recent oncology clinical trials, retrospective data analysis following completion of clinical trials can result in development of gene signatures that can be predictive of patient response.^1–5^ However, traditional methods to identify actionable biomarkers are time consuming, expensive, mostly prognostic, and often fail to provide actionable insights.

To identify patterns within high dimensional multiomics data, artificial intelligence algorithms have been developed to assist in understanding basic patterns within the data.^6–10^ At the more advanced level, complex neural networks could, in theory, identify non-linear complex relationships of clinical and biologic information that can predict treatment benefit, which cannot be achieved by classical methods but could be made feasible by the assistance of advanced AI models.^11^ For this reason, we sought to develop a Large ClinicoGenomic foundation Model (LCGM) that can leverage the complexity of high dimensional data (clinical and multiomic) to predict treatment benefit. If the model can learn complex biological relationships in the foundation model, then the LCGM should be able to identify patterns that predict treatment benefit in new datasets.

Our technology utilizes a deep learning architecture and training schema to predict the benefit of a clinical trial intervention, which we define as the benefit of a therapeutic intervention (experimental arm) compared to the standard of care (control arm). The name of our LCGM is Clinical trials Uncovering Real Efficacy Artificial Intelligence, or CURE AI^TM^. CURE AI predicts which patients benefit more from an investigational therapy than standard of care at the individual patient level based on their baseline clinicogenomic profile. Identifying patient populations more likely to benefit from treatment, a process known as predictive enrichment, can contribute to the development of robust and reliable patient enrichment strategies.^12–13^ CURE AI enables predictive enrichment though enhanced classification of true responders and true non-responders to therapy. Accurately defining eligibility criteria is a key part of ensuring that patients are offered therapy that they will benefit from, which is an important consideration for new drug approval.^14–16^

CURE AI tackles two major bottlenecks in clinical drug development. The first problem is data scarcity (limited patient numbers and data missingness) combined with the high dimensionality of clinical data. Clinical trial data often presents challenges due to limited patient numbers that cannot be easily pooled into larger databases as well as a vast amount of data points per patient (>1,000,000 measurements in phase 3 trials with omics data).^17–18^ Using a novel and proprietary deep learning architecture and training schema, we developed an approach to not only identify the most important clinicogenomic variables but to learn the connections and patterns between these variables on a much larger scale using our LCGM trained on a large corpus of patient data. The second challenge is sorting out the differences in prognostic and predictive markers. Current methods in clinical trials often find correlations between variables but are unable to assess all available clinical and multiomic data to pinpoint patterns within that data that explain why drugs benefit certain patient groups but not others.^19^ Inability to accurately identify predictive factors can result in misinterpretation of trial outcomes, with immortal time bias being a well-known example.^20^ For instance, a patient’s functional status (prognostic factor) might be mistakenly interpreted to drive their health outcome to a new drug, leaving the true predictive factors undiscovered. The difficulty lies in predicting the benefit that an individual would receive from both the investigational therapy and the standard of care, which allows for a prediction of the magnitude of excess benefit derived from the investigational therapy. We need to be able to predict response to both trial therapies to design and interpret the most personalized and effective clinical trials.

To achieve this, a perfect clinical trial dataset would contain every patient’s response to both trial therapies (randomizing each patient to both trial arms), which would allow measurement of the real benefit of the investigational therapy over the existing standard of care at the individual patient level. Since it is not possible to administer different therapies to a patient in parallel, we developed CURE AI to accurately predict this difference for each patient based on accurate comparisons of their clinical and genomic characteristics to similar patients in what can be thought of as an advanced implementation of causal inference.^21–22^ In this seminal manuscript, we validate the premise of CURE AI to guide patient therapy selection in two completed advanced non-small cell lung cancer randomized clinical trials using one as validation set and the other as held-out test set. We show how CURE AI can guide clinical trial interpretation and design by AI-guided prediction of therapeutic benefit.

## Methods

### CURE AI Development

CURE AI is a deep learning technology developed by Numenos which consists of two main parts: (1) a base neural architecture foundation model that was trained on a large cohort of clinicogenomic datasets in a proprietary self-supervised learning schema with multiple loss function^23^ and (2) a proprietary training process to finetune CURE AI on indication-specific clinical data for the specific downstream task of predicting treatment benefit using k-fold cross-validation (CV)^24^ and out-of-fold (OOF) predictions. CURE AI contains a foundation model trained on a large cohort of patients with diverse pathologies including all major cancer types.

The CURE AI LCGM was finetuned on progression-free survival (PFS)-outcome stratified random selection of four-fifths of the patients on the OAK trial for whom RNA sequencing was available. The model was then used to predict benefit on the held-out one-fifth of the validation patients. This process was iterated five total times (to define the features that predict treatment benefit on the entire included OAK trial dataset) to create the finetuned CURE Lung Cancer^TM^ model. A schematic of the CURE AI training model is shown in Figure 1.

**Figure 1.**
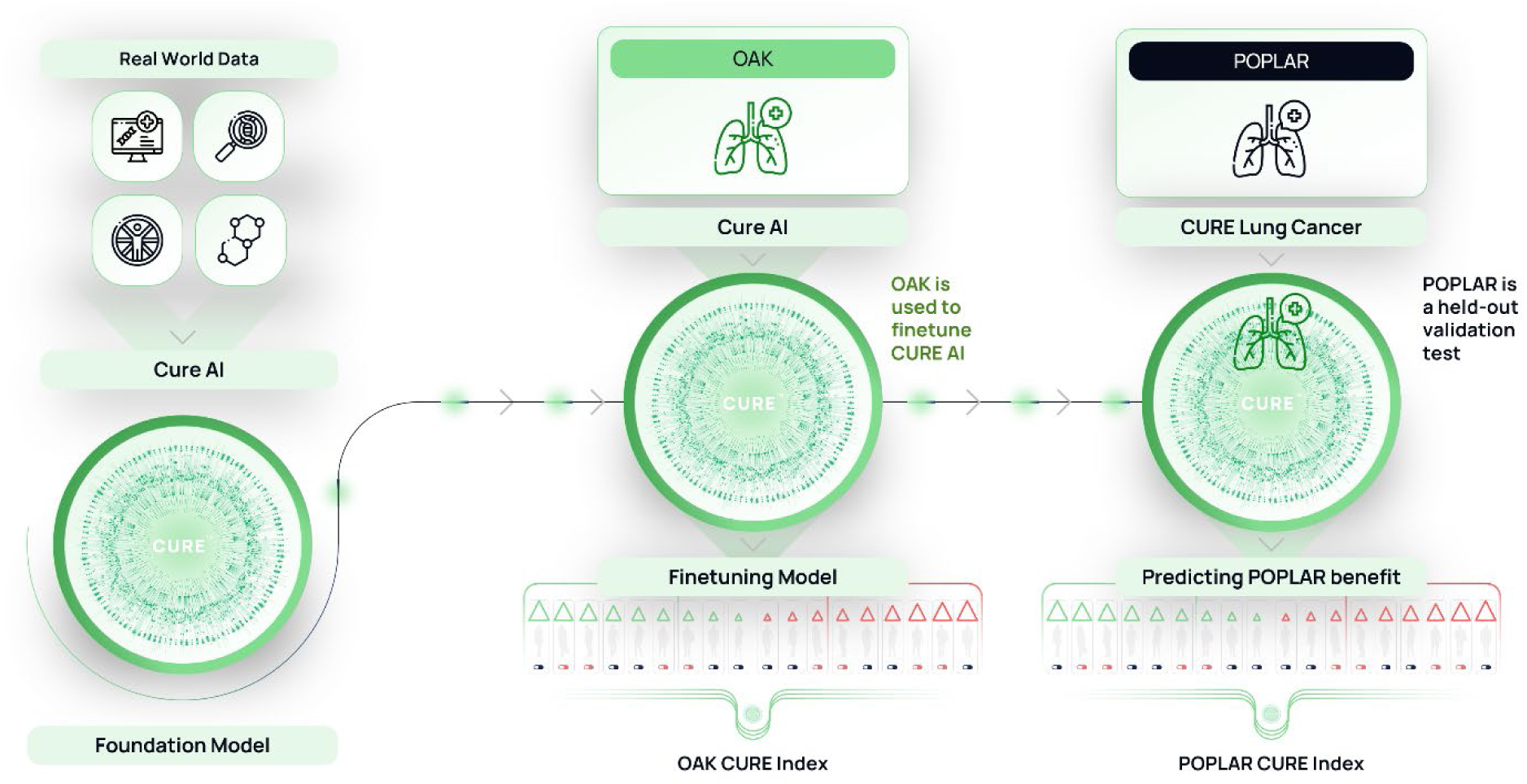
Outline of CURE AI Development, Finetuning, and Validation Testing. The CURE AI foundation model was trained on vast clinical and multiomic datasets to learn and understand the interplay between clinical and multiomic features. CURE AI was applied to the OAK clinical trial to finetune the model for training (finetuning CURE AI into CURE Lung Cancer using conditional average treatment effect modeling) and cross-validation (testing how well CURE AI predicts outcomes on OAK). CURE Lung Cancer was then validation tested on the completely held-out POPLAR clinical trial dataset. The CURE Index is created as an output from the models, from which we define CURE AI-informed simulated trial enrollment.

### CURE Score^TM^ and CURE Index^TM^ Evaluation of Individual Patient Benefit

CURE AI calculates a counterfactual outcome for each patient to predict how each patient would have responded to the trial therapy that they did not receive.^25^ This individualized patient benefit prediction is called the CURE Score. The estimate of benefit of investigational therapy over standard of care in a specific trial is based on finetuning CURE AI with Conditional Average Treatment Effect (CATE) modeling.^26^ The CURE Score is calculated by assessing the difference of the predicted treatment outcomes. A positive score favors investigational therapy, and a negative score favors standard of care.

The ordered ranking of each patient within a clinical trial by their CURE Score is called the CURE Index. The CURE Index is used to define subgroups of patients. We create a Cox-proportional hazard model, with the treatment arm being the only covariate for each subgroup. Because of randomization, benefit prediction is equally distributed between the trial arms, allowing a balanced dataset to train the Cox-proportional hazard model and measure treatment effect using the hazard ratio for the treatment covariate for that subgroup. The log-rank test is used to generate the p-value for therapeutic benefit. Thus, treatment effect is based on the hazard ratio and significance is based on the p-value.

### The CURE Curve

We developed the CURE Curve as the aggregated p-value plot of all patients included in the order of the CURE Index. The CURE Curve presents CURE AI results and allows for straightforward identification of the patients who benefit more from either the investigational therapy or the standard of care. We enroll each patient based on their order in the CURE Index, starting from the highest CURE Score. For each additional patient enrolled (who received either atezolizumab or docetaxel), a p-value is calculated to assess whether we can reject the null hypothesis that there are no differences in outcome between the treatment groups. If we observe significance in CURE Index-defined subsets (p < 0.05), then CURE AI successfully identified patient characteristics that predict better response to atezolizumab.

In order to compare the CURE Curve to a random baseline, we simulated trial enrollment of patients based on random enrollment (rather than CURE index-defined enrollment, we used random ranking). A p-value is calculated for the random enrollment, which is performed 1000 times. The p-value line for random patient enrollment is presented with its mean value as well as 1 standard deviation on each side of the mean. At every point where the CURE Curve p-value is above the upper confidence interval of the random selection line (ie: the p-value has a lower absolute value for CURE Indexed patients) CURE AI is significantly outperforming random trial enrollment.

Because the CURE Score is based on baseline information, CURE Scores are directly comparable for patients on similar clinical trials. We can therefore compare groups of patients across different trials (comparing OAK patients to POPLAR patients), based on each patient’s CURE Score.

The CURE Curve generates “natural subgroups” of patients with similar predicted benefit values. We selected three representative points along the OAK and POPLAR trial CURE Curves that represent two possible inclusion thresholds generated by CURE AI (2 thresholds plus the entire dataset for each trial). For the OAK trial, we selected the top 35% and 52% of CURE Score-defined patients. The trial percentiles for POPLAR trial patients with the same CURE Scores corresponded to the top 44% and 88% of patients.

### CURE Permutation Test^TM^

We developed a permutation test as an additional assessment of confidence in CURE AI. The permutation test involves randomizing the outcome of each patient so that the treatment outcome is no longer correctly associated with the same patient. This is done by randomizing PFS independently within each trial arm, so that the distribution of outcomes per trial arm remains the same. Permutated data from OAK and POPLAR were independently analyzed by CURE AI as described above. If the permutated data leads to loss of significance (loss of relationship between clinical variables and outcome), then we are assured of more confidence in our model.

### Kaplan-Meier Estimation of Progression Free Survival

We plot the Kaplan-Meier estimates for the probability of progression-free survival for three representative points along the CURE Curve for both the OAK and POPLAR trials. We utilized PFS as the outcome because the OAK and POPLAR trials both failed to meet this endpoint.

Additionally, PFS is most directly related to cancer treatment effect and therefore less prone for bias, whereas overall survival can be affected by factors unrelated to cancer treatment or progression. Kaplan-Meier plots were generated using Python. The number of patients at risk (alive and evaluable; non-censored) at each timepoint are shown for the non-permutated plots.

### Clinical Trial Datasets

The phase 3 OAK and phase 2 POPLAR advanced non-small cell lung cancer randomized prospective clinical trials both tested the investigational therapy atezolizumab compared to docetaxel in previously treated advanced non-small cell lung cancer.^3,4^ We restricted our analysis to patients who had available RNA-sequencing data, which led to the inclusion of 699 patients on the OAK trial and 193 patients on the POPLAR trial. Tabular clinical trial data were integrated into CURE AI as training and testing variables.

Pre-processing of Bulk RNA Sequencing data:

Raw read counts and transcripts per million (TPM) were utilized for sample normalization. These data integrate into CURE AI as training and testing variables.

## Results

CURE AI predicts a benefit score for each patient, which allows enrolled patients to be ranked from high to low predicted benefit to investigational therapy compared to standard of care (Figure 1). This order of patients is called the CURE Index. We then enroll each patient by the CURE Index order and calculate a p-value for each additional enrolled patient, which generates the CURE Curve. We developed the CURE Curve as a visual representation of the performance of CURE AI in predicting clinical trial treatment benefit.

The CURE Curves and associated Kaplan-Meier estimates for the OAK trial^3^ are demonstrated in Figure 2. The OAK trial represents a validation of the performance of the CURE AI foundation model because the benefit prediction was finetuned on OAK using out-of-fold benefit prediction derived only from baseline OAK patient profiles (see Figure 1). Practically, this means that CURE AI is a high-performing foundation model as patients who benefit from atezolizumab more than docetaxel could be identified through analysis of a single clinical trial.

**Figure 2.**
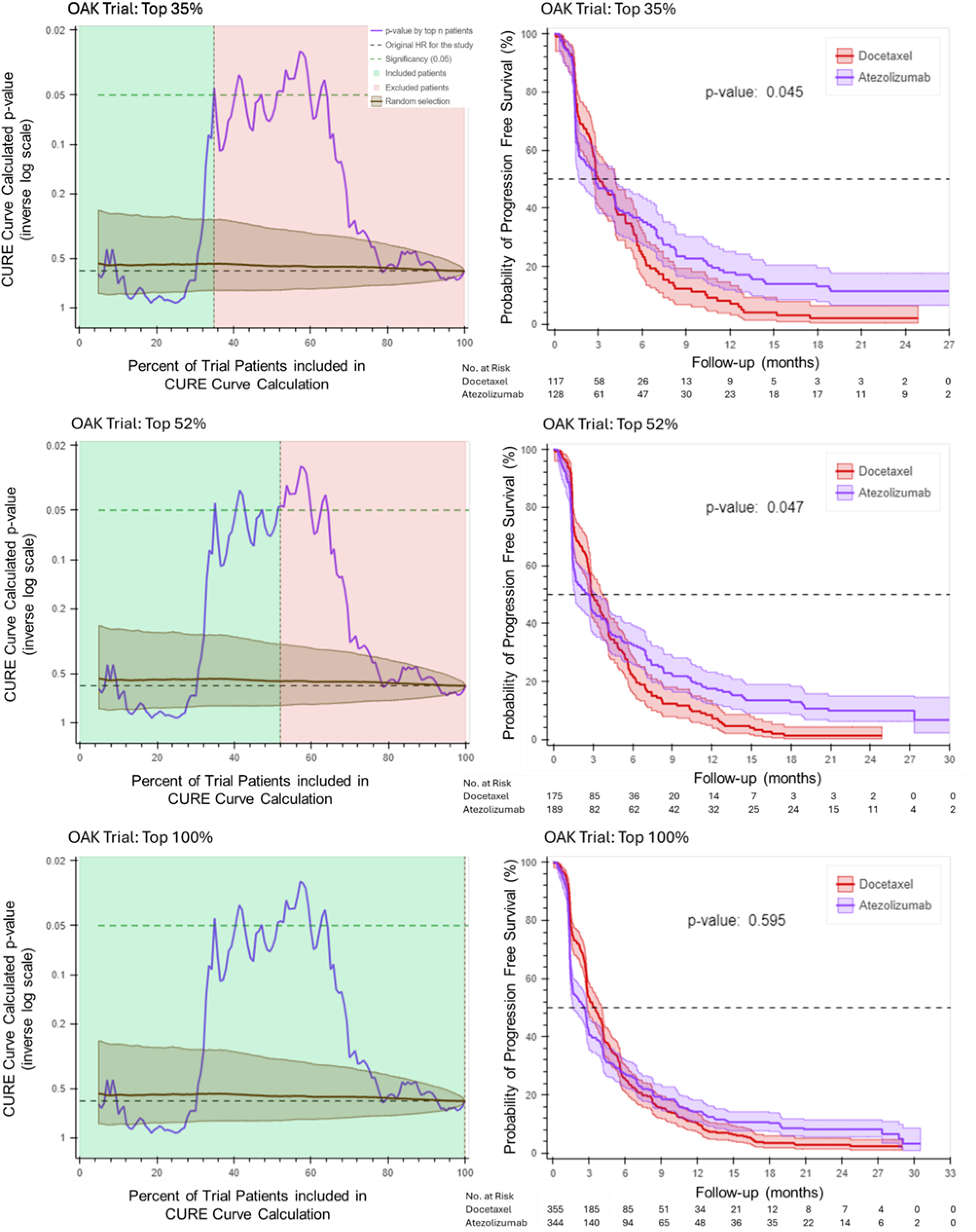
CURE Curves and Kaplan-Meier Estimates for the OAK Trial. In the OAK trial, the CURE Curve shows that patients enrolled up to around the 65^th^ percentile (where the p-value line begins to descend) are more likely to benefit from atezolizumab than docetaxel. When the CURE Curve begins to descend, patients who have more predicted benefit to docetaxel are included, which results in loss of trial significance in the total trial population as the standard of care arm outperforms atezolizumab in the >65^th^ percentile subgroup. Top Panel. The top 35% of CURE Index defined patients were selected. At this threshold, the CURE Curve reaches significance (p < 0.05), with the corresponding Kaplan-Meier estimation shown on the right panel. This CURE Index is equivalent to the top 44% of patients on the POPLAR trial. Middle Panel. The top 52% of CURE Index defined patients were selected. At this threshold, the CURE Curve remains above significance (p < 0.05), with the corresponding Kaplan-Meier estimation shown on the right panel. This CURE Index is identical to the top 88% of patients on the POPLAR trial. Bottom Panel. The entire included trial population is shown with the corresponding Kaplan-Meier estimation shown on the right panel (p = 0. 595). For each Kaplan-Meier estimate, the number of patients at risk (alive and evaluable) at each timepoint is shown.

Two OAK trial CURE Curve thresholds were selected for Kaplan-Meier estimation. These two thresholds represent the first point that the CURE Curve reaches significance (top 35% of OAK patients, corresponding to the top 44% of POPLAR patients) and the point on the POPLAR trial that is the most inclusive for patients who experience more benefit from atezolizumab relative to docetaxel (the top 88% of patients on POPLAR, which corresponds to the top 52% of the OAK CURE Index). These two thresholds represent the same CURE Score, meaning that they have the same predicted benefit to atezolizumab relative to docetaxel.

The point where the CURE Curve p-value first reaches significance is upon enrollment of the top 35% of the CURE Index patients (Figure 2, top panel). The Kaplan-Meier estimates for progression-free survival calculation for the 35% threshold is shown in Figure 2, top right panel (p = 0.045 indicating significant benefit to atezolizumab in this group). Figure 2 (middle panels) demonstrate the CURE Curve and associated Kaplan-Meier estimates for the CURE Curve threshold of 52% (p = 0.047). Both of the CURE Curve thresholds significantly outperform the entire training set (which is shown in Figure 2, bottom panels) which has a p-value of 0.595. At every point along the CURE Curve up to when the p-value line begins to descend, that patient cohort (all patients enrolled from the highest CURE Index and to that point) has more benefit to atezolizumab than docetaxel. The point where the CURE index p-value begins to consistently decrease (around 65% on the OAK CURE Curve) represents the threshold where patients benefit more from docetaxel than from atezolizumab.

To address concerns of overfitting, we compared CURE AI performance to a random recruitment of patients. The random selection line on the CURE Curve includes an average of 1000 iterations of simulated randomized trial enrollment of the same number of included patients to that point along the CURE Curve. A p-value is calculated for this patient group (the cohort receiving atezolizumab compared to the cohort receiving docetaxel) for each enrollment iteration, and the p-value is averaged. The standard deviation for random patient enrollment p-value is shown on the CURE Curve. At every point where the CURE Curve p-value line is above the upper confidence interval of the random selection line, CURE AI significantly outperforms random trial enrollment.

We hypothesized that if a trial is well balanced, CURE AI should not generate an imbalance in trial arms. Indeed, we found that the CURE Curve-defined groups remain well balanced in both patient number and clinical features. Table 1 demonstrates that some of the major variables associated with clinical response are not imbalanced due to CURE patient selection for the OAK trial including gender, histology, TMB^27^, and PD-L1 status. These data demonstrate that CURE AI can effectively predict benefit to therapy on the OAK trial while keeping baseline characteristics balanced and despite significant data missingness (PD-L1 was only defined for about half of patients, for example).

**Table 1.**
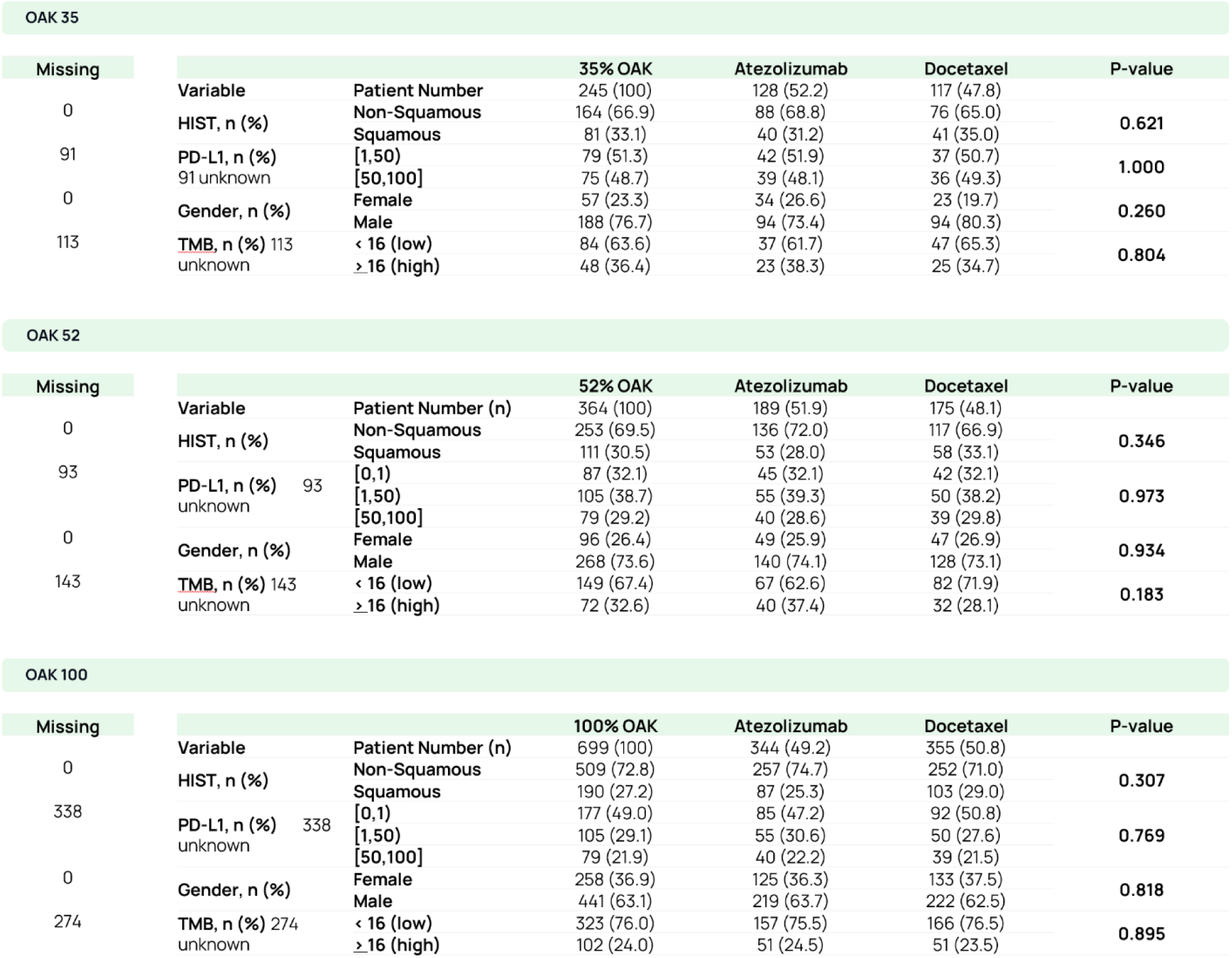
OAK Trial Patient Demographics by CURE Curve Threshold (top 35% of patients, top table; top 52% of patients, middle table; all included patients, bottom table). Number of patients with an unknown variable is listed in the left column. These data demonstrate that CURE Lung Cancer patient group demographics are not significantly altered compared to all included patients for the OAK trial.

We deliberately only included the OAK trial in the finetuning process to showcase two points: (1) how CURE AI performs on a real clinical trial when used for both training and cross-validation and (2) to assess the performance on a smaller and completely held-out trial (POPLAR), which we describe next. CURE AI, now finetuned on OAK non-small cell lung cancer data, is termed CURE Lung Cancer.

CURE Lung Cancer demonstrated direct generalizability on a held-out independent clinical trial dataset, the POPLAR trial.^4^ The POPLAR trial has a similar patient population and treatment arms (atezolizumab vs. docetaxel) as the OAK trial. We utilized CURE Lung Cancer to calculate the CURE Curve for the POPLAR clinical trial. The CURE Curves and Kaplan-Meier estimates for the POPLAR trial are demonstrated in Figure 3. The top panel demonstrates the top 44% of the CURE Index patients (corresponding to the top 35% of patients on the OAK trial, which represents the same CURE Score). We find that the PFS is significant with a p-value of 0.021 (compared to 0.211 on the entire patient cohort; Figure 3, bottom panel).

**Figure 3.**
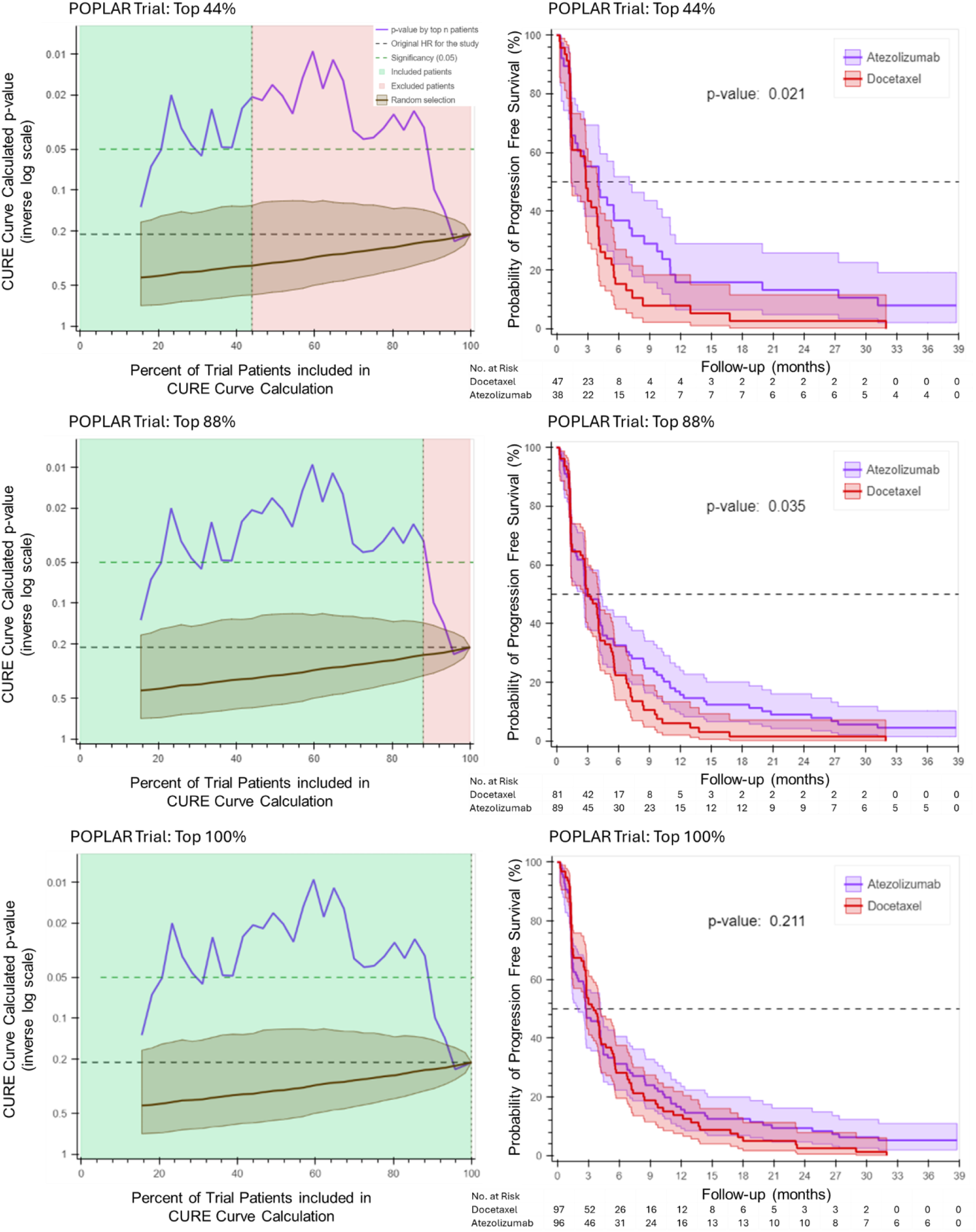
CURE Lung Cancer Curves and Kaplan-Meier Estimates for the POPLAR Trial. In the POPLAR trial, the CURE Curve shows that patients enrolled up to around the 88^th^ percentile (where the p-value line begins to descend) are more likely to benefit from atezolizumab than docetaxel. When the CURE Curve begins to descend, patients who have more predicted benefit to docetaxel are included, which results in loss of trial significance in the total trial population as the standard of care arm outperforms atezolizumab in the >88^th^ percentile subgroup. Note that the POPLAR trial was analyzed on CURE Lung Cancer (CURE AI that has been finetuned on the OAK trial). Top Panel. The top 44% of CURE Index defined patients were selected. At this threshold, the CURE Curve passes significance (p < 0.05), with the corresponding Kaplan-Meier estimation shown on the right panel. Middle Panel. The top 88% of CURE Index defined patients were selected. At this threshold, the CURE Curve remains above significance (p < 0.05), with the corresponding Kaplan-Meier estimation shown on the right panel. Bottom Panel. The entire included trial population is shown with the corresponding Kaplan-Meier estimation shown on the right panel (p = 0. 211). For each Kaplan-Meier estimate, the number of patients at risk (alive and evaluable) at each timepoint is shown.

Interestingly, the top 52% of patients on the OAK trial corresponded to the top 88% of patients on the POPLAR trial, based on the same CURE Score cutoffs. This means that the patient group on the POPLAR trial contained more patients who we predict would have more benefit to atezolizumab. We find that on POPLAR, if the trial included the top 88% of patients by CURE Index ranking, the trial would have had a positive PFS endpoint (p = 0.035).

Since the POPLAR trial was well balanced, CURE AI should not lead to an imbalance in trial arms. Indeed, we found that the CURE Curve-defined groups remain well balanced in both patient number and clinical features. Table 2 demonstrates that major variables associated with clinical response are not imbalanced due to CURE patient selection for the POPLAR trial. These data demonstrate that CURE Lung Cancer can effectively predict benefit to therapy for the POPLAR trial, which is a completely independent test of CURE AI training and finetuning process.

**Table 2.**
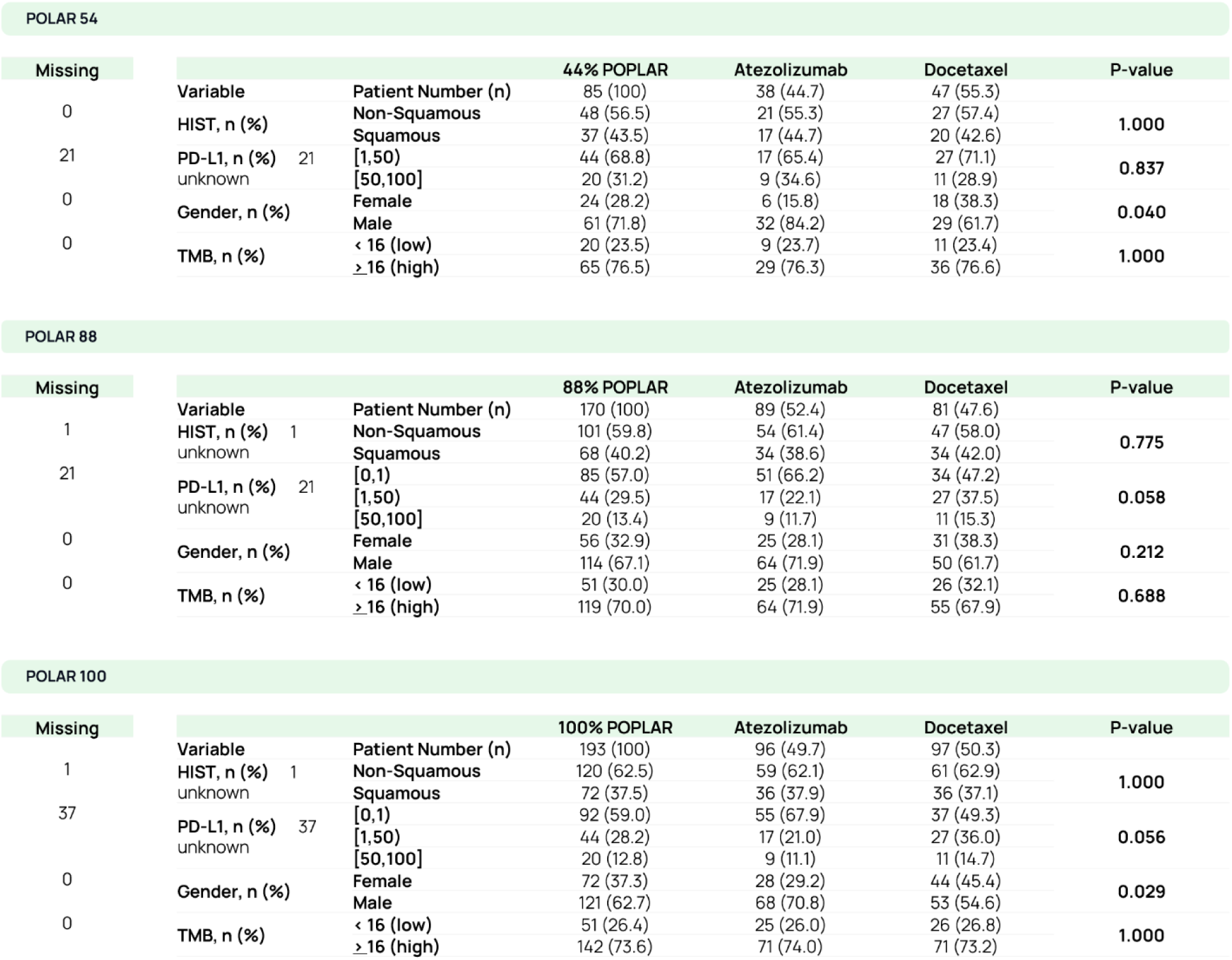
POPLAR Trial Patient Demographics by CURE Curve Threshold (top 44% of patients, top table; top 88% of patients, middle table; all included patients, bottom table). Number of patients with an unknown variable is listed in the left column. These data demonstrate that the CURE Lung Cancer patient group demographics are not significantly altered compared to all included patients for the POPLAR trial.

Since the design of CURE AI intrinsically limits the risk of confounding by validation testing on held-out trial data, and we saw no signs of data overfitting or bias within our models, we have high confidence in the performance of CURE AI. Indeed, our finetuned model generalized to the POPLAR trial. However, as an additional test to affirm our confidence in the model, we developed the CURE Permutation test to assess for intrinsic bias within the model finetuning schema. The permutation test involves randomizing the outcome of each patient so that treatment outcomes are no longer correctly associated with the same patient’s clinicogenomic information. Within each trial arm, PFS is randomized so that the distribution of outcomes per trial arm remains the same, but the association of PFS for a particular patient is incorrect. This should result in loss of signal for the model training, as the clinicogenomic relationships that are predictive for patient benefit are no longer accurate, so the model should not be able to learn an association since the correct association no longer exists.

Permutated patient outcome data from OAK and POPLAR were independently assessed by CURE AI, as described in the schema from Figure 1. Figure 4 demonstrates the CURE Permutation test on the OAK Trial. We find that there is a loss of significance across the entire dataset, as the CURE Curve no longer outperforms the randomized patient selection. At the top 35% of the CURE Index (Figure 4, top panel), the p-value is insignificant at p = 0.391, compared to p = 0.045 on the CURE AI model for OAK. At the top 52% of the CURE Index (Figure 4, bottom panel), the p-value is insignificant at p = 0.703, compared to p = 0.047 on the CURE AI model for OAK. Figure 5 demonstrates the CURE Permutation test on the POPLAR Trial. We find that there is a loss of significance across the entire dataset, as the CURE Curve no longer outperforms the randomized patient selection. At the top 44% of the CURE Index (Figure 5, top panel), the p-value is insignificant at p = 0.629, compared to p = 0.021 on the CURE AI model for POPLAR. At the top 88% of the CURE Index (Figure 5, bottom panel), the p-value is insignificant at p = 0.371, compared to p = 0.035 on the CURE AI model for POPLAR. The results demonstrate that CURE AI is not prone to generating false associations between data and outcomes.

**Figure 4.**
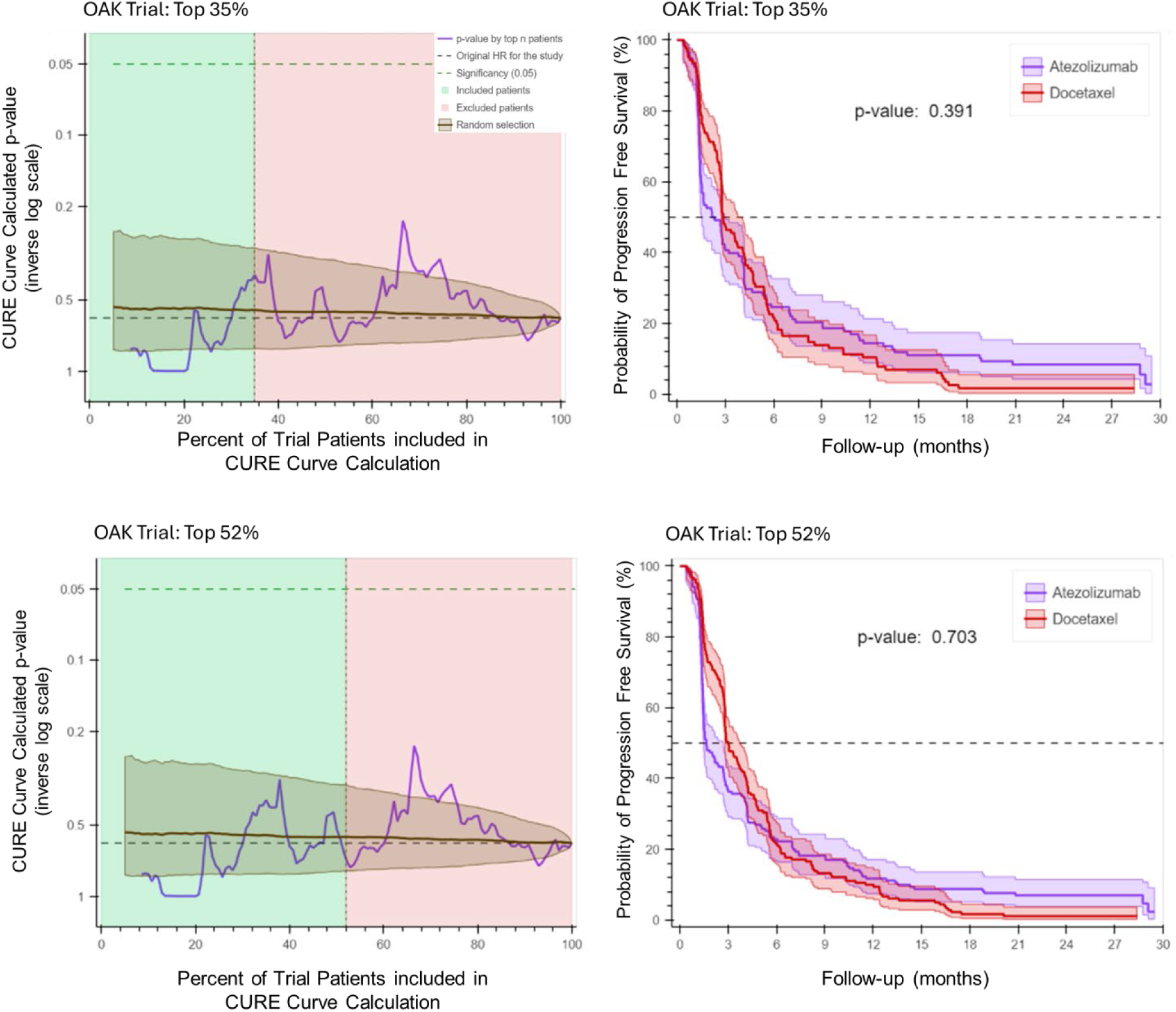
CURE Permutation Test and Kaplan-Meier Estimates for the OAK Trial. Using the CURE Permutation test, when PFS is randomized within each arm of the OAK trial, CURE AI does not identify any significant associations between clinicogenomic variables and patient outcome. The top 35% and top 52% of CURE Index defined patients during permutation testing are demonstrated. The CURE Curve is not significantly different than the random selection enrollment (p > 0.05 at all points), with the corresponding Kaplan-Meier estimates shown on the right panel. This loss of significance is a test of confidence in the CURE AI model.

**Figure 5.**
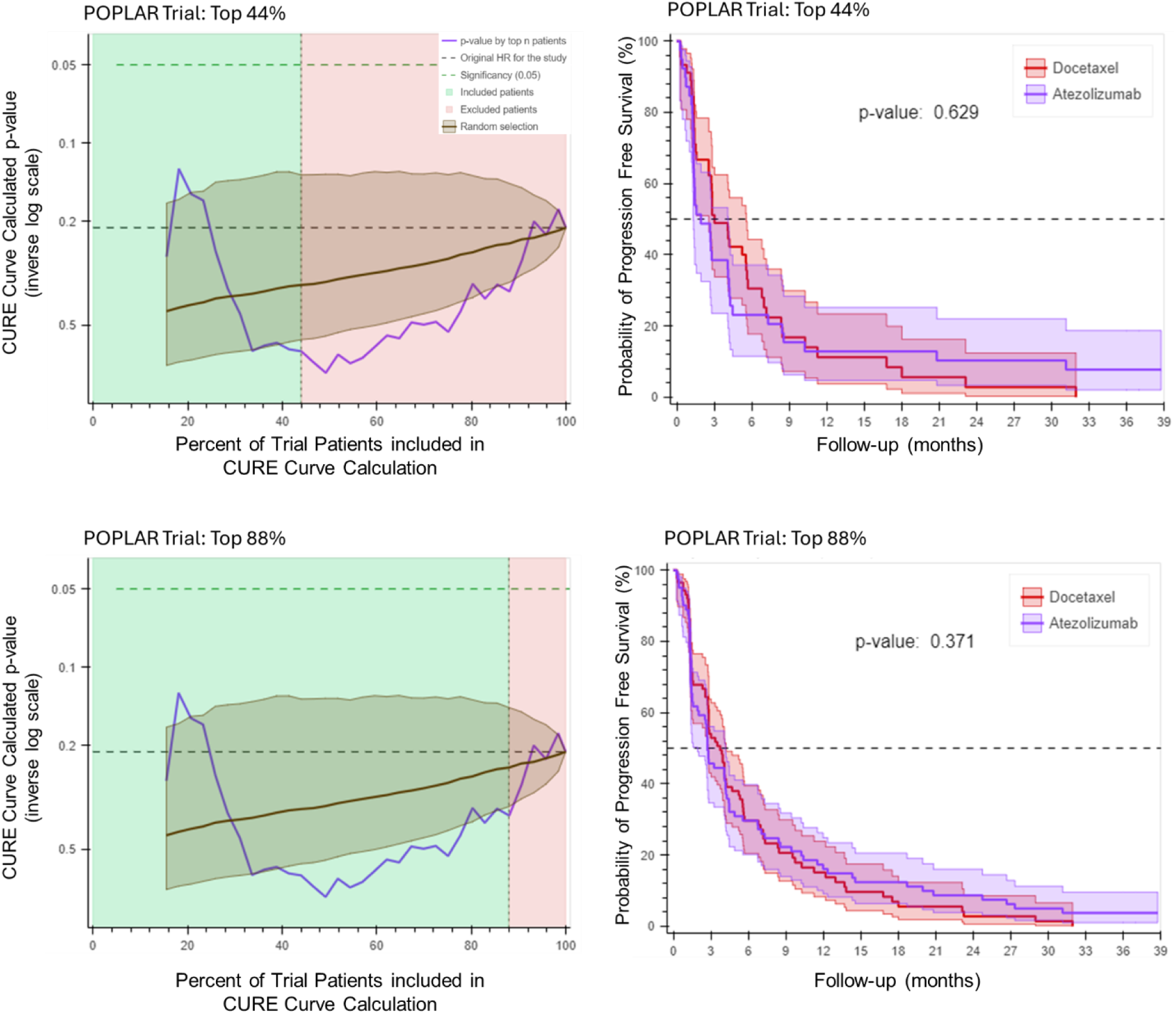
CURE Permutation Test and Kaplan-Meier Estimates for the POPLAR Trial. Using the CURE Permutation test, when PFS is randomized within each arm of the POPLAR trial, CURE AI does not identify any significant associations between clinicogenomic variables and patient outcome. The top 44% and top 88% of CURE Index defined patients during permutation testing are demonstrated. The CURE Curve is not significantly different than the random selection enrollment (p > 0.05 at all points), with the corresponding Kaplan-Meier estimates shown on the right panel. This loss of significance is a test of confidence in the CURE AI model.

## Discussion

The CURE AI foundation model is an innovative example of how complex neural network architectures can be applied to clinical trials to increase the likelihood that patients will receive a drug for which they will benefit. We independently validated the performance of CURE AI on two clinical trials and showed that CURE AI can predict patient benefit while being blinded to patient outcome. This is the first demonstration that an LCGM can proactively characterize patients by predicted benefit to an investigational therapy.

While further validation is required, based on the presented evidence it is attractive to speculate that LCGMs will cause a shift in the way clinical trials are approached through enhancement of enrollment criteria that enhances patient selection for treatment benefit. With improved enrollment criteria, we should observe a Will Rogers effect, where the clinical outcomes of all patients screened for a trial will improve. This is because patients who are eligible for enrollment will be enriched for benefit to investigational therapy, and patients who are screened ineligible will avoid receiving an investigational therapy that is predicted to be inferior to the existing therapeutic option. For example, on POPLAR, the top 88% of patients should be enrolled and randomized. However, the patients who are predicted to benefit more from standard of care could instead be offered standard of care alone or an alternative clinical trial. In practice, CURE AI could be used to understand which actionable clinical and genetic factors define benefit from a phase 2 trial, which would refine eligibility criteria for a phase 3 trial. Another exciting use case is to adapt ongoing clinical trials based on interim analyses, in which patient eligibility criteria could be modified during the trial to optimize for therapeutic benefit.

CURE AI has the potential to improve clinical trial design, analysis, and interpretation. In the future, it is possible that LCGMs will enable ‘virtual control arms’ for clinical trials, as standard control arms could raise concerns for not offering treatment with high predicted benefit over standard of care, based on existing data. Regulatory bodies will need to collaborate with academic, biotechnology, and patient advocacy leaders to ensure safe, transparent, and straightforward implementation of technologies like CURE AI, as the clinical translation of transformative patient treatments could be rapidly expedited with the assistance of these technologies.

In summary, we have developed a causal LCGM that can predict and accurately measure individualized patient benefit of investigational therapy compared to an existing standard of care. The development of CURE AI enables a shift from simple outcomes (such as treatment response) to individualized continuous benefit measurement. We are optimistic that special implementations of AI, such as CURE AI for clinical trials, will demonstrate consistent successes and lead to a revolution in healthcare including a major step toward more individualized medicine. CURE AI can be applied to a variety of scientific and clinical uses including adaptive clinical trials, toxicity prediction, treatment response prediction, and understanding of drug resistance and response mechanisms.

## Data Availability

All data generated or analyzed during this study that are available are included in this published article. The code underlying all CURE AI models is a proprietary trade secret. CURE AI models are currently available to access by commercial entities and academic collaborations or for non-profit organizations for clinical trial design and analysis through Numenos. The models are not currently available for independent public research use.

## Authors’ contributions

AW and VF conceived and developed CURE AI. AW, NP and VF designed the trial analyses in this study. AW, VF, and NP analyzed and interpreted the trial analyses in this study. DF, ZT, JC, and BJ contributed to the supervision and project strategy. All authors contributed to writing and reviewing the manuscript. All authors have read and approved the final manuscript.

## Ethics approval and consent to participate

Not applicable.

## Patient consent for publication

Not applicable.

